## Supplemental Material for "Evidence for Influenza and RSV interaction from 10 years of enhanced surveillance in Nha Trang, Vietnam, a modelling study"

**Affiliations**:
[1] Centre for Mathematical Modeling of Infectious Disease, London School of Hygiene and Tropical Medicine, UK
[2] Department of Pediatrics, Nagasaki University Hospital, Nagasaki University, Nagasaki 852-8102, Japan
[3] Department of Pediatric Infectious Diseases, Institute of Tropical Medicine, Nagasaki University, Nagasaki 852-8523, Japan
[4]Statistics, Modelling and Economics Department, UKHSA
[5]National Institute of Hygiene and Epidemiology, Hanoi, Vietnam

* These authors have contributed equally

**Estimated influenza attack rate** 2
**Correlation** 3
**Model equations** 3
**R0 equations** 5
**Susceptibility to RSV** 5
**Susceptibility to Influenza** 6
**Parallel tempering** **6**
**Attack Rates** 9
**Sensitivity to severity of dual infected cases** 10
**Prior Sensitivity** 10
**References** 11

#### 1. Estimated influenza attack rate

Assuming no interaction (in susceptibility to or severity of dual infections), we calculated the required annual influenza infection attack rate in order to achieve the observed number of dual infections (equations 1-3). Using a negative binomial likelihood with Brent optimization we estimated the RSV reporting rate that would correspond to the maximum likelihood of observing the reported weekly number of dual infections. We then used this estimate of the reporting rate to calculate the annual RSV population attack rate required in order to observe this many dual cases. The confidence intervals for the attack rate were calculated using the Hessian matrix from the optimisation.

$$I_{Dual} \simeq I_{RSV} * P_{Influenza} \quad (1)$$

$$P_{Influenza} \simeq I_{Influenza} * 1/\gamma_{Influenza} / \upsilon_{Influenza} \quad (2)$$

$$AR_{Influenza} \simeq I_{Influenza} / \upsilon_{Influenza} \quad (3)$$

With parameters: Incidence of reported cases (I), Prevalence of Infection (P), Duration of Infection( $1/\gamma$ , 3.8 days - see main text Table 1) and estimated reporting rate **(**$\upsilon$).

We estimated that in order to achieve the weekly reported number of dual infections given no interaction, we would require an annual influenza attack rate of 4.4 (3.4 -6.5) in ages 0-1 and 1.3 (0.9 - 3.0) in ages 2-4.

#### 2. Correlation

Figure S1 shows a scatter plot between the weekly Influenza and RSV cases.

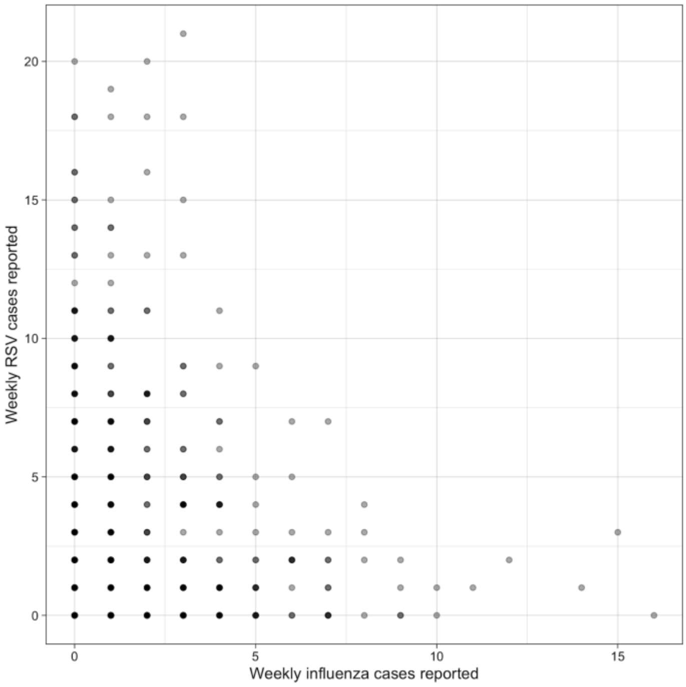


**Figure S1: Reported cases**. Scatter plot of weekly influenza and RSV cases reported through the enhanced surveillance study in less than 5 year olds over the whole time period.

#### 3. Model equations

Full model equations are shown below. Each compartment includes the state for both RSV and influenza, with the first letter indicating the state for RSV, and the second for influenza. E.g. $SS_i$ is shorthand for $S_{RSV,i}S_{INF,i}$. Subscripts used are “INF” for influenza and “RSV”

$$\lambda_{INF,i} = \sum_{j=1}^{5} \beta_{INF}\alpha_{ij}I_{INF,j}$$

$$\lambda_{RSV,i} = \sum_{j=1}^{5} \beta_{RSV}\alpha_{ij}I_{RSV,j}$$

$$\frac{dSS_i}{dt} = -\tau_i\lambda_{RSV_i}SS_i - \lambda_{INF_i}SS_i - \epsilon_{INF} - \epsilon_{RSV}$$

$$\frac{dIS_i}{dt} = \tau_i\lambda_{RSV_i}SS_i - (1-\sigma)\lambda_{INF_i}IS_i - \gamma_{RSV}IS_i + \epsilon_{RSV}$$

$$\frac{dPS_i}{dt} = \gamma_{RSV} IS_i - \rho PS_i - (1-\sigma)\lambda_{INF_i} PS_i$$

$$\frac{dRS_i}{dt} = \rho PS_i - \lambda_{INF_i} RS_i$$

$$\frac{dSI_i}{dt} = \lambda_{INF_i} SS_i - (1-\sigma)\tau_i \lambda_{RSV_i} SI_i - \gamma_{INF} SI_i + \epsilon_{INF}$$

$$\frac{dII_i}{dt} = (1-\sigma)\lambda_{INF_i} IS_i + (1-\sigma)\tau_i \lambda_{RSV_i} SI_i - \gamma_{INF} II_i - \gamma_{RSV} II_i$$

$$\frac{dPI_i}{dt} = \lambda PS_i - \gamma_{INF} PI_i + \gamma_{RSV} II_i + \lambda_{INF_i} RS_i$$

$$\frac{dSP_i}{dt} = \gamma_{INF} SI_i - \rho SP_i - (1-\sigma)\tau_i \lambda_{RSV_i} SP_i$$

$$\frac{dIP_i}{dt} = (1-\sigma)\tau_i \lambda_{RSV} SP_i - \gamma_{RSV} IP_i + \gamma_{INF} II_i + \tau_i \lambda_{RSV_i} SR_i$$

$$\frac{dSR_i}{dt} = \rho SP_i - \tau_i \lambda_{RSV_i} SR_i$$

$$\frac{dRR_i}{dt} = \gamma_{RSV} IP_i + \gamma_{INF} PI_i$$

Where:

$\lambda_{i,j}$ - force of infection between age groups I and J

$\beta$- transmission rate

$\alpha_{ij}$- contact rate between group I and j

$\tau_i$ - age group susceptibility to RSV

$\sigma$ - level of cross-protection

$\gamma$ - rate of recovery

$\rho$ - rate of loss of cross-protection

$\epsilon$ - introduction rate from external sources

#### 4. $R_0$ equations

The R0's were calculated as the dominant eigenvalue of the matrix

$$-T\Sigma^{-1}$$

Where $T$ is the transmission matrix, describing new infections, and$\Sigma$ is the transition matrix, describing other changes in state. This method is described in full in Diekmann *et al (2009)*[1]

$$T_{INF} = \begin{bmatrix} \beta_{INF} * \alpha_{i,j} & \cdots & \beta_{INF} * \alpha_{i,j} \\ \vdots & \ddots & \vdots \\ \beta_{INF} * \alpha_{i,j} & \cdots & \beta_{INF} * \alpha_{i,j} \end{bmatrix} T_{RSV} = \begin{bmatrix} \tau_j * \beta_{RSV} * \alpha_{i,j} & \cdots & \tau_i * \beta_{RSV} * \alpha_{i,j} \\ \vdots & \ddots & \vdots \\ \tau_j * \beta_{RSV} * \alpha_{i,j} & \cdots & \tau_j * \beta_{RSV} * \alpha_{i,j} \end{bmatrix}$$

$$\Sigma_{INF} = \begin{vmatrix} \gamma_{INF} & 0 & 0 & 0 & 0 \\ 0 & \gamma_{INF} & 0 & 0 & 0 \\ 0 & 0 & \gamma_{INF} & 0 & 0 \\ 0 & 0 & 0 & \gamma_{INF} & 0 \\ 0 & 0 & 0 & 0 & \gamma_{INF} \end{vmatrix} \Sigma_{\mathrm{RSV}} = \begin{vmatrix} \gamma_{RSV} & 0 & 0 & 0 & 0 \\ 0 & \gamma_{RSV} & 0 & 0 & 0 \\ 0 & 0 & \gamma_{RSV} & 0 & 0 \\ 0 & 0 & 0 & \gamma_{RSV} & 0 \\ 0 & 0 & 0 & 0 & \gamma_{RSV} \end{vmatrix}$$

The posterior estimates led to $R_0$s of 1.07 (95%CrI 1.06-1.1) and 1.24 (95%CrI 1.23 - 1.26) for influenza and RSV respectively.

#### 5. Susceptibility to RSV

We used a longitudinal study by Hendersen et a. (1979) to determine age-susceptibility to RSV infection. They estimated that at 1st exposure 98.4% of children became infected, at second exposure 74.5% of children became infected and at 3rd exposure 65.4% of children became infected[2]. As most children are infected by 24 months of age, we used the susceptibility estimates for the age groups: ages 0-1 = 100% susceptible, ages 2-4 = 75% susceptible, ages 5 and over = 65% susceptible[3].

#### 6. Susceptibility to Influenza

Influenza susceptibility each season (s) is determined by the parameter $\eta_s$ using the inverse density of an exponential distribution at each age group (where ages 0-1 is age group zero, up to ages 65+ at age group four). Example susceptibility profiles are shown in Figure S2.

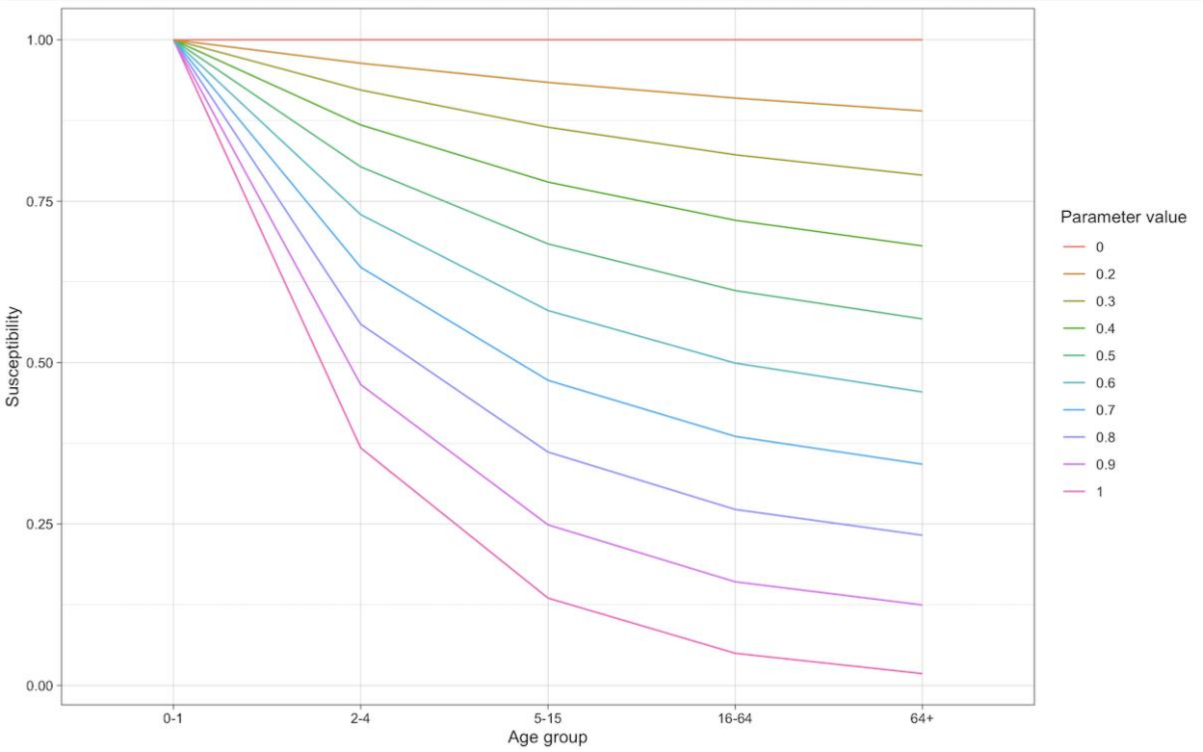


***Figure S2: Susceptibility.*** *Susceptibility to influenza by age group for different parameter values.*

#### 7. Parallel tempering

Our parallel tempering algorithm was implemented in R and we used Amazon Web Services (AWS) to run it. We proposed swaps with the next temperature chains every 5 iterations. We ran the parallel tempering algorithm using a covariance matrix to propose parameters. We removed 250000 iterations as burn in, followed by 200000 more samples, and assessed convergence using the Geweke statistic in the null chain (Chain with temperature 1). This calculates the difference between the two sample means of the first 10 and last 50% of the chain, divided by its estimated standard error, resulting in a Z score. Note however that due to the large number of parameters (44), the multiple modes and the swapping between chains as a result of the parallel tempering the Geweke statistic is not an ideal measure of convergence in this situation. Despite this, all key parameters (transmission rates, interaction parameters, dual detection rate) had a Z score within the 95% confidence interval and overall over 80% of parameters fell within a 99% confidence interval. Figure S3 shows one of the traces for a sample of parameters, thinned to 1 in 10. Figure S4 shows the final posterior densities of each parameter, and Figure S5 describes the distribution. We also ran a second set of chains starting at different parameter values, which converged on the same parameter spaces.

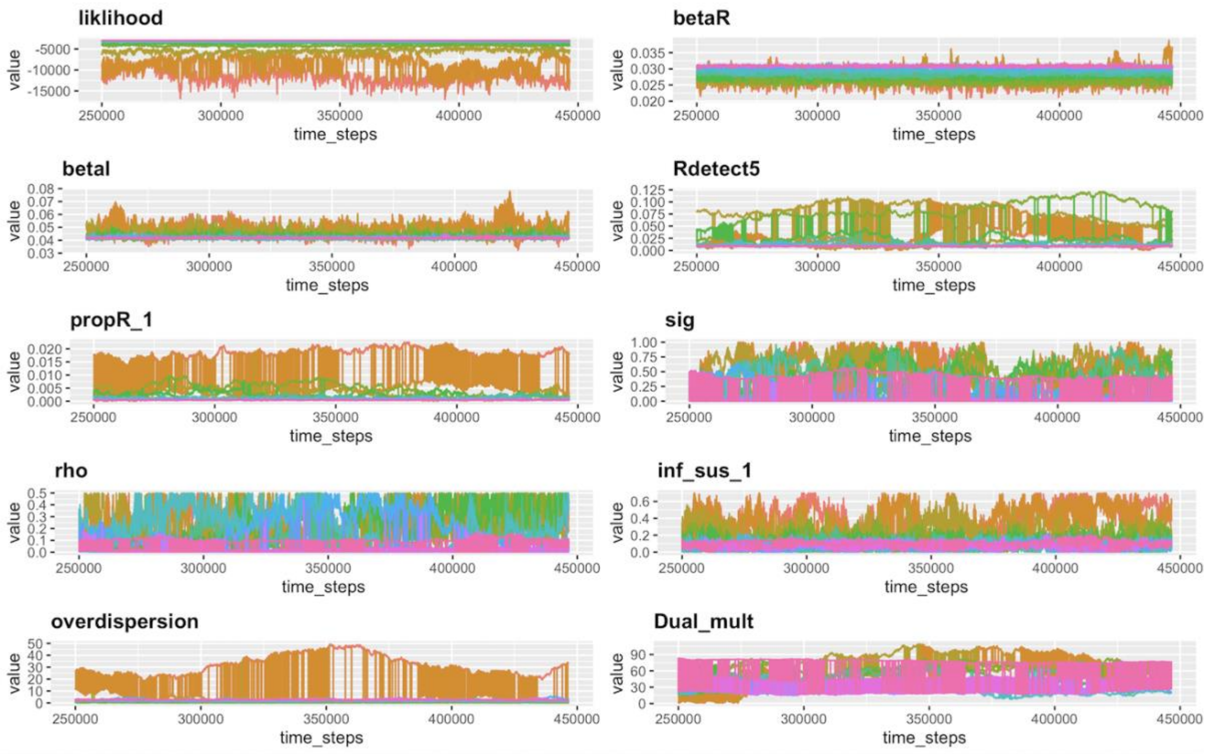


***Figure S3: Parallel Tempering Trace.*** *Sample trace from parallel tempering, showing a subsection of parameters. Each colour is a chain at a different temperature, where the main chain is pink and the chain at the highest temperatures is red. The chains are thinned to 1:10.*

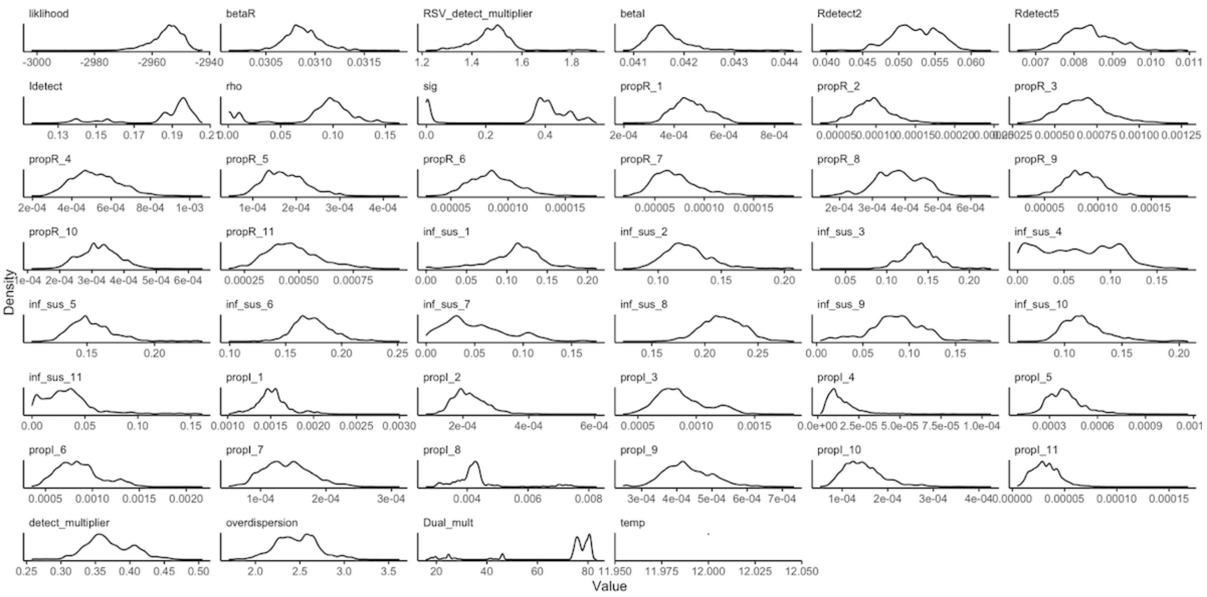


***Figure S4: Posterior Density.*** *Density of fitted parameters from the final sample.*

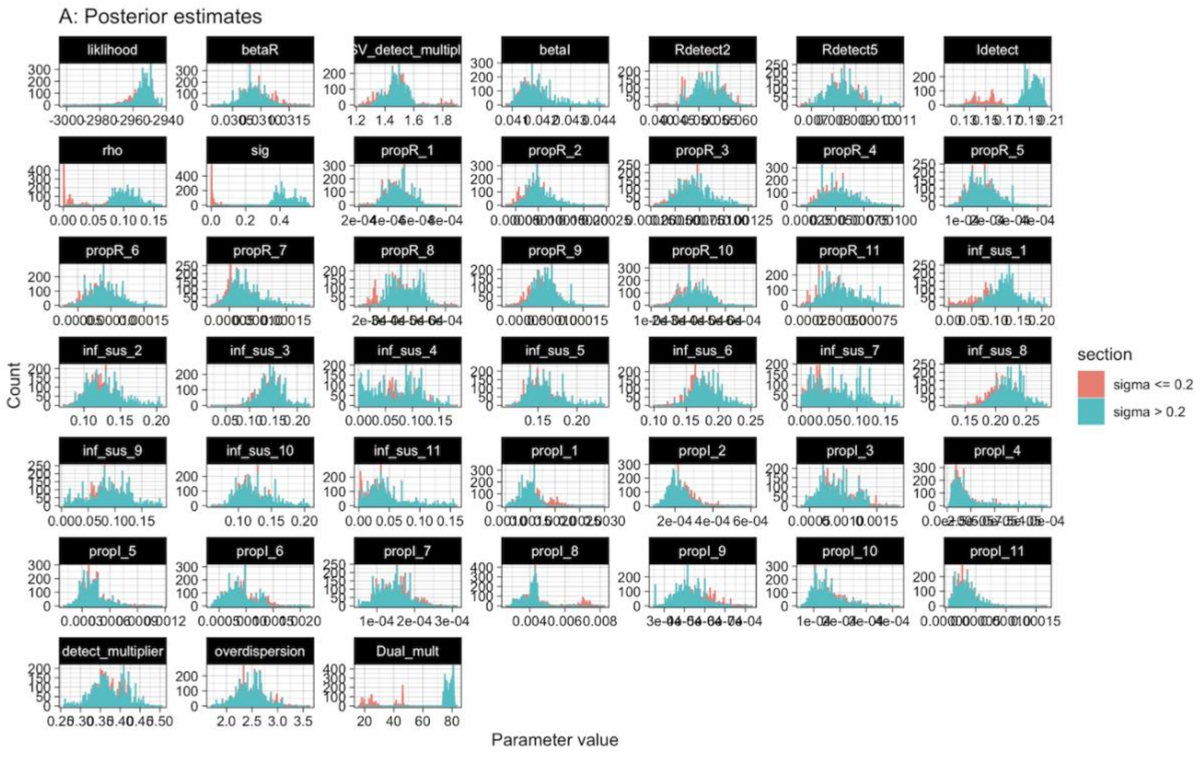


***Figure S5: Posterior parameter estimates, split by the value of sigma (interaction parameter)***

### 8. Attack Rates

Figure S5 shows the Attack Rates for each virus, season and age group, as well as the susceptibility to influenza at the start of the season by age group, calculated from 50 posterior samples.

We estimated a seasonal attack rate ranging from 24% to 41% for RSV and 1% to 15% for Influenza. For RSV, the attack rate was lowest in the oldest age group of 65+, whereas for influenza the lowest attack rates were in the youngest age group of 0-1 years old. Susceptibility to influenza at the start of the season was high, with all age groups in all years being over 87% susceptible to infection with the circulating strain.

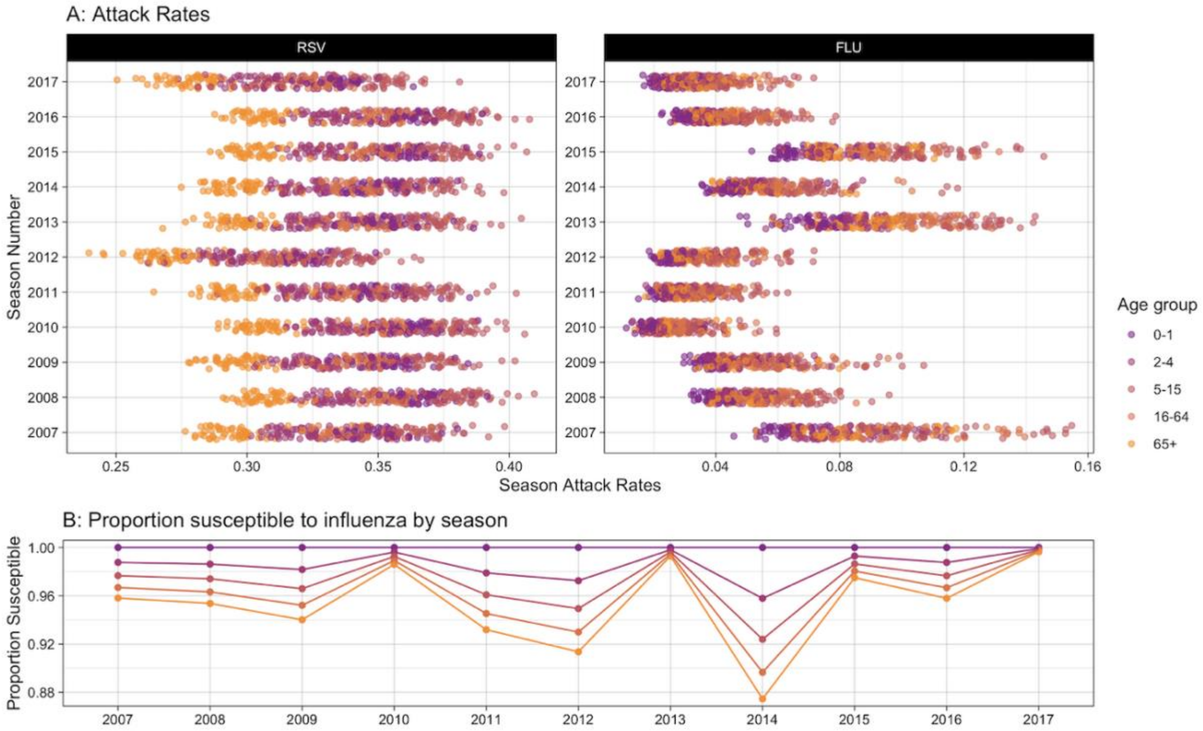


***Figure S6: Modelled Output.*** *A) Season attack rates for influenza and RSV by age group. B) Proportion susceptible to influenza at the beginning of the season for each year by age group, using the median value of the posterior samples. Each year the susceptibility of each age group is defined by one parameter in an exponential function, see supplement for details.*

### 9. Sensitivity to severity of dual infected cases

We tested the assumption of the severity of dual infected cases, by rerunning the fit without the parameter that multiplied the proportion of RSV detected to give a new dual infection detection rate. Instead the dual infections had the same reporting rate as for RSV. This set of chains were run for 100000 iterations and 50000 was discarded as burnin, and then the remaining samples were thinned to 1 in 10. Cross-protection estimates overlapped with estimates of the ‘no interaction’ mode in the main model, with the posterior for interaction at 0.008 (95%Ci 0.00 - 0.04) compared to 0.004 (95%CI 0.000 - 0.046).

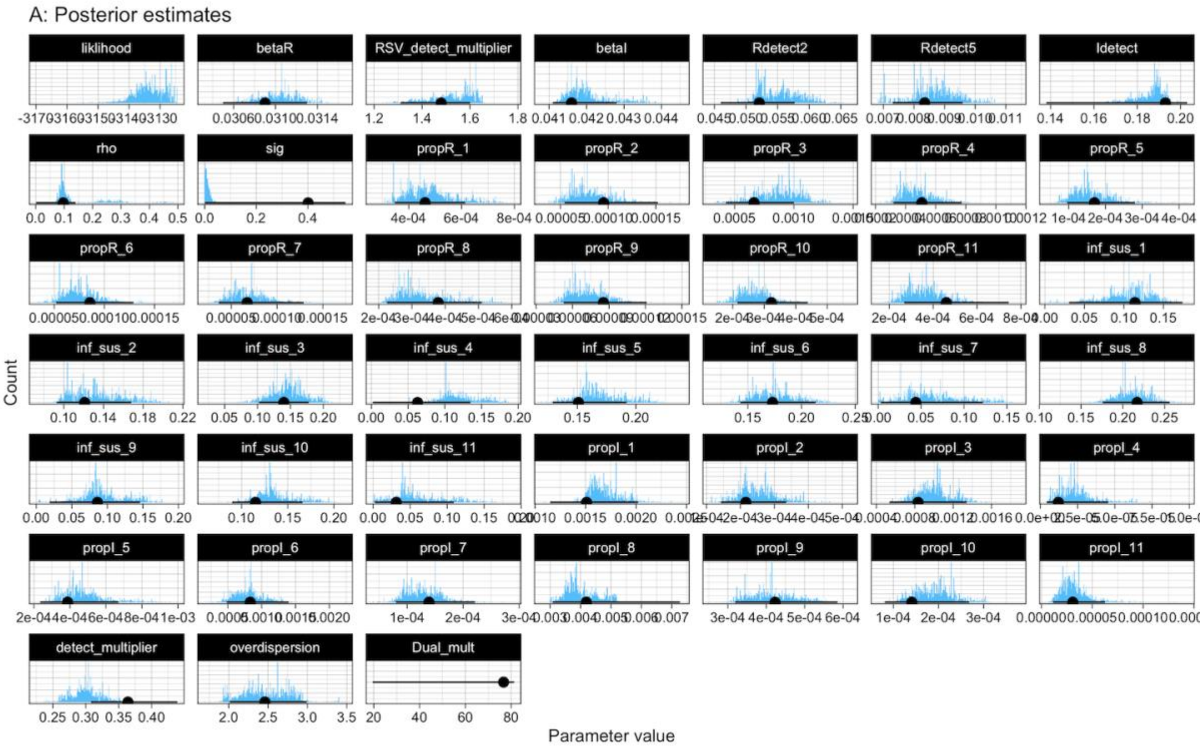


***Figure S6: Parameter Density.*** *Density of fitted parameters from the final sample. Black lines show the median and 95%CI for the main model run.*

### 10. Prior Sensitivity

As a sensitivity analysis, we reran the model fit with a prior for a high strength of interaction (normal distribution, mean = 0.8, standard deviation = 0.15). This is due to the existing evidence of cross-protection. Figure S6 shows the posterior estimates for the parameters. This set of chains were run for 100000 iterations and 25000 was discarded as burnin, and then the remaining samples were thinned to 1 in 10. Cross-protection estimates overlapped with estimates of the ‘moderate interaction’ mode in the main model, with the posterior for interaction at 0.22 (95%Ci 0.13 - 0.47) compared to 0.41 (95% 0.36 - 0.54) and the duration of cross-protection at 5.2 days (95%CI  3.1 -10) compared to 10.0 days (95%CI 7.1 -12.8 days).

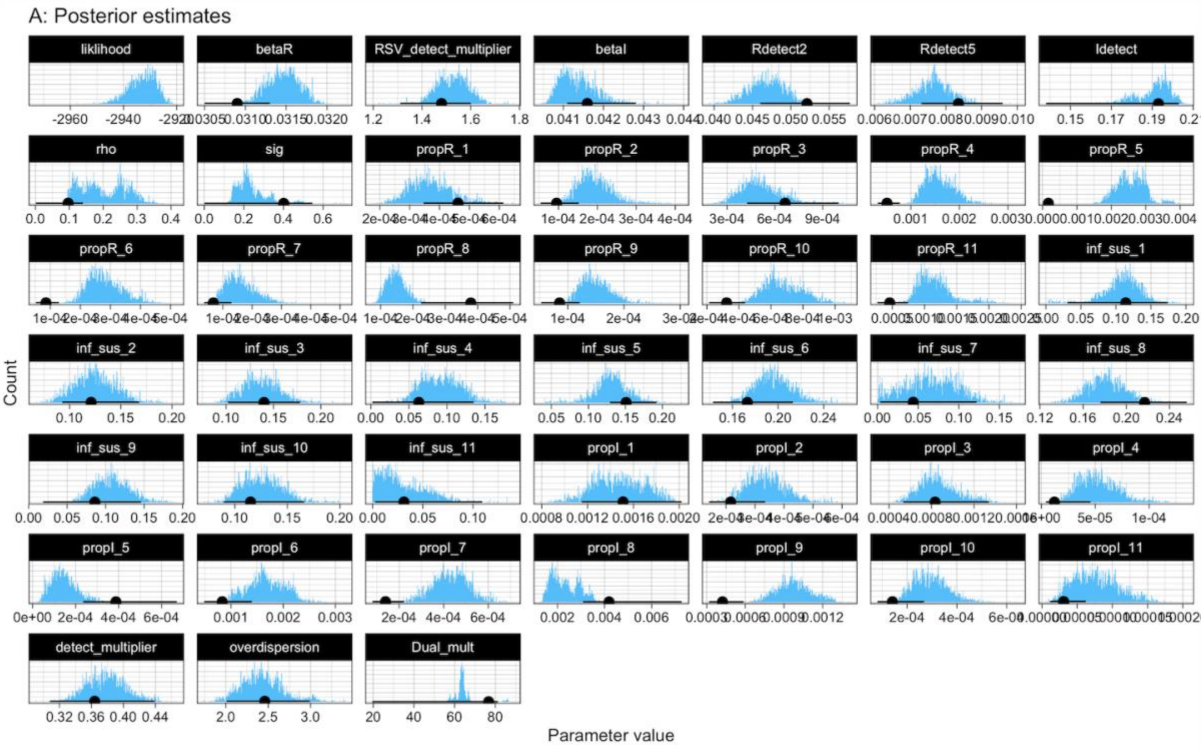


***Figure S6: Parameter Density with interaction prior.*** *Density of fitted parameters from the model with a prior for strong cross-protection.*
